# The Silent Signal: Unmasking Myocardial Ischemia in a Resting Heartbeat with Machine Learning

**DOI:** 10.1101/2025.11.01.25339306

**Authors:** Basheer Abdullah Marzoog, Philipp Kopylov

## Abstract

**Background:** Ischemic heart disease (IHD) remains the leading cause of morbidity and mortality worldwide, imposing a staggering burden on healthcare systems and societies.

**Aims:** To assess the diagnostic capabilities of the single lead electrocardiography in the diagnosis of IHD using the machine learning model.

**Materials and methods:** A prospective, non-randomized, minimally invasive, single-center, case-control study enrolled male and female participants aged ≥40 years. All participants underwent resting single-lead electrocardiography (SLECG) and pulse wave recording using a portable Cardio-Qvark® device, and stress computed tomography myocardial perfusion imaging with vasodilation test. Based on coronary computed tomography perfusion (CTP) results, 80 participants were stratified into two groups: Group 1 with stress-induced myocardial perfusion defect (n = 31) and Group 2 without stress-induced myocardial perfusion defect (n = 49). Statistical processing carried out using the R programming language v4.2, Python V.3, Statistica 12 programme. (StatSoft, Inc. (2014). STATISTICA (data analysis software system), version 12. www.statsoft.com.), and SPSS IBM version 28. P considered statistically significant at <0.05.

**Results:** The best model performance in the diagnosis of IHD was Random Forest model. The model showed a diagnostic accuracy based on the parameters of the SLECG, AUC 0.988 [95 % confidence interval (CI); 0.967-1.000], Sensitivity 0.871 [95 % CI; 0.739-0.971], Specificity 0.959 [ 95 % CI; 0.894-1.000].

**Conclusion:** This study demonstrates that machine learning analysis of resting single-lead ECG signals, acquired via the portable Cardio-Qvark® device, achieves near-perfect accuracy (AUC 0.988) in diagnosing ischemic heart disease validated against myocardial perfusion imaging.

## Introduction

Ischemic heart disease (IHD) remains the leading cause of morbidity and mortality worldwide, imposing a staggering burden on healthcare systems and societies [1]. Characterized by an imbalance between myocardial oxygen supply and demand, most commonly due to atherosclerotic coronary artery disease (CAD), IHD manifests across a spectrum from silent ischemia and stable angina to acute coronary syndromes and heart failure [2]. Early and accurate detection is paramount, as timely intervention can significantly alter disease progression, prevent catastrophic events like myocardial infarction, and improve long-term outcomes [3–5]. Despite decades of advancement, however, the diagnostic pathway for IHD continues to present significant challenges, often balancing the need for accuracy against considerations of accessibility, cost, invasiveness, and patient risk.

Conventional diagnostic strategies rely heavily on a combination of clinical assessment, resting electrocardiography (ECG), and non-invasive stress testing. While the standard 12-lead ECG is ubiquitous, inexpensive, and non-invasive, its diagnostic sensitivity for detecting ischemia, particularly in stable patients without active symptoms or in those with single-vessel disease, is notoriously limited [6]. Resting ECG findings can be normal in a substantial proportion of individuals with significant CAD. Consequently, functional stress testing – such as exercise ECG, stress echocardiography, or myocardial perfusion imaging (MPI) with single-photon emission computed tomography (SPECT) or positron emission tomography (PET) – is frequently employed to provoke ischemia [7]. While more sensitive, these tests often require specialized facilities, are time-consuming, relatively expensive, involve radiation exposure (in the case of nuclear MPI), and carry inherent risks associated with physical or pharmacological stress agents, limiting their suitability for widespread screening or use in certain patient populations. Coronary computed tomography angiography (CCTA) offers excellent anatomical detail but also involves radiation and iodinated contrast, and its functional significance for detected stenoses often requires further testing. Invasive coronary angiography, the historical gold standard for defining coronary anatomy, is reserved for higher-risk patients due to its invasive nature, cost, and associated procedural risks.

This diagnostic conundrum has fueled intense interest in developing more accessible, rapid, and highly accurate non-invasive tools for IHD detection. The proliferation of portable and wearable cardiac monitoring devices offers a promising avenue. These technologies enable the acquisition of physiological data, including electrocardiographic signals, outside traditional clinical settings, potentially facilitating earlier detection and continuous monitoring [8]. Single-lead ECG (SLECG) devices, in particular, represent a paradigm of simplicity and portability, capable of capturing essential cardiac electrical activity with minimal setup. However, the true diagnostic potential of these devices often remains unrealized. The raw data they provide, typically limited to heart rate and basic rhythm analysis, lacks the sophistication needed for robust ischemia detection. Translating the rich information embedded within the ECG waveform into a reliable diagnostic tool for IHD necessitates advanced analytical methods capable of discerning subtle, complex patterns indicative of underlying myocardial ischemia that escape conventional interpretation.

Machine learning (ML), a subset of artificial intelligence, has emerged as a transformative force in medical diagnostics. By learning complex patterns and relationships from large datasets without being explicitly programmed for every rule, ML algorithms offer the potential to extract novel insights from existing data sources, such as the ECG [9]. Applied to ECG analysis, ML models can integrate and interpret a multitude of waveform features – including morphologies, intervals, amplitudes, and variability metrics – far beyond the capabilities of traditional human reading or simple automated algorithms. This capability holds immense promise for enhancing the diagnostic yield of simple, readily available tools like SLECG, potentially elevating them from basic rhythm monitors to sophisticated ischemia detectors. Previous research has explored ML for ECG analysis, often using standard 12-lead systems, demonstrating feasibility in detecting conditions like atrial fibrillation or hyperkalemia, and showing encouraging, though often variable, results for IHD detection. However, the application of rigorously validated ML models specifically to resting SLECG data acquired via portable devices for the purpose of diagnosing IHD, using stress MPI as a robust reference standard, represents a significant and relatively underexplored frontier.

This study, therefore, aimed to bridge this critical gap by developing and validating a machine learning model specifically designed to diagnose IHD non-invasively using resting SLECG parameters acquired with a portable device. We utilized the Cardio-Qvark® system, a CE-marked device capable of recording both SLECG and photoplethysmogram (PPG)-derived pulse waves. The primary objective was to leverage ML algorithms to analyze the rich feature set extracted from these resting SLECG recordings and determine their diagnostic accuracy in identifying patients with stress-induced myocardial perfusion defects, as defined by the current clinical gold standard of stress computed tomography perfusion (CTP) imaging. We hypothesized that advanced computational analysis of resting SLECG signals could uncover latent patterns highly predictive of underlying myocardial ischemia, enabling the development of a highly accurate, point-of-care screening tool that overcomes many limitations of current diagnostic approaches. By employing rigorous model development techniques, including stratified cross-validation and bootstrap resampling for confidence intervals, and focusing on interpretable ML models to identify key predictive features, this research seeks to provide a robust foundation for a novel, accessible, and potentially transformative approach to IHD detection. The ultimate goal is to contribute a practical tool that can facilitate earlier diagnosis, optimize resource utilization by reducing unnecessary referrals for advanced testing in low-risk individuals, and improve patient outcomes through timely intervention, particularly in settings where access to sophisticated cardiac diagnostics is limited.

## Materials and methods

### Study design

A prospective, non-randomized, minimally invasive, single-center, case-control study enrolled male and female participants aged ≥40 years, reflecting the marked increase in coronary heart disease risk beyond this age. Recruitment occurred at Sechenov University’s University Clinical Hospital No. 1 between October 27, 2023, and October 28, 2024. Patient data with pathology were sourced from the Department of Cardiology, while healthy participant data were collected retrospectively. The study adhered to Good Clinical Practice standards and the Declaration of Helsinki, was registered on ClinicalTrials.gov (NCT06181799), and received approval from the Sechenov University Ethics Committee (Approval number 19-23; October 26, 2023). All participants provided written informed consent for study participation and publication of results, including associated figures.

Based on coronary computed tomography perfusion (CTP) results, 80 participants were stratified into two groups: Group 1 with stress-induced myocardial perfusion defect (n = 31) and Group 2 without stress-induced myocardial perfusion defect (n = 49). The sample size (n = 80) was determined through power analyses for related means and Pearson correlation using SPSS software.

### Data collection

The study assessed both continuous and categorical variables. Continuous parameters included age, body weight, height, resting pulse, systolic blood pressure (SBP), diastolic blood pressure (DBP), echocardiographic ejection fraction (EF%), estimated vessel age, creatinine, and estimated glomerular filtration rate (eGFR; calculated using the CKD-EPI 2021 equation). Categorical variables encompassed demographics (gender, obesity stage, smoking status), clinical conditions (concomitant diseases, coronary artery involvement, presence of hemodynamically significant stenosis >70%), and perfusion status (pre- and post-stress ATP perfusion defects).

Participant selection criteria were defined according to the specific aim and goal of the study. This structured assessment of variables facilitated the analysis in alignment with the study’s objectives.

1. All participants underwent resting single-lead electrocardiography (SLECG) and pulse wave recording using a portable Cardio-Qvark® device (Moscow, Russia) [10]. Recordings were acquired continuously over a 3-minute period. The resulting SLECG and pulse wave data were analyzed using machine learning models developed by the Sechenov University research team [10, 11]. Comprehensive documentation of the specific Cardio-Qvark® parameters utilized in this study is provided in the referenced publication and the clinical trial protocol (NCT06181799) [12].
2. Prior to coronary artery multidetector computed tomography (MDCT) with myocardial perfusion assessment, all participants underwent serum creatinine measurement to calculate glomerular filtration rate (GFR) via the CKD-EPI formula. Eligibility required a GFR ≥30 mL/min/1.73 m², consistent with contrast safety protocols [13–16]. Radial vein catheterization was then performed for administration of Iohexol contrast agent (Omnipaque®; 50 mL; GE Healthcare) and sodium adenosine triphosphate (ATP) to pharmacologically induce myocardial ischemia through heart rate elevation.

Imaging was conducted using a Canon Aquilion One Genesis scanner (Roszdravnadzor No. RZN 2021/16161) with a 640-slice protocol: initial 0.5 mm native slices followed by contrast-enhanced perfusion imaging at rest and 20 minutes post-ATP infusion. Triphosadeninum (ATP; Ellara®; LP-004667) was diluted in saline (3 ampoules [10 mg/mL] + 17 mL NaCl 0.9%) and administered as a weight-adjusted slow IV bolus (300 μg/kg over 2 minutes). Cardiothoracic radiologists interpreting CT perfusion data remained blinded to stress ECG results throughout the analysis.

Image evaluation employed the American Heart Association’s 16-segment model, with layered assessment of basal, mid-ventricular, and apical segments. Short-axis projection analysis first identified confounding artifacts, followed by systematic perfusion defect detection. Segments were mapped to coronary territories:

LAD: Segments 1,2,7,8,13,14

LCx: Segments 5,6,11,12,16

RCA: Segments 3,4,9,10,15

Perfusion defects were identified visually as regions of reduced X-ray attenuation during the ATP-induced stress phase, indicating ischemia. Automated analysis using Vitrea Advanced software calculated transmural perfusion ratios (TPR) and generated rest/stress polar maps. Perfusion status was classified by a five-tier color scale:

Blue (TPR 2.5–0.99): Normal

Green (0.99–0.97): Mild abnormality

Yellow (0.97–0.94): Moderate hypoperfusion Orange (0.94–0.60): Severe hypoperfusion Red (0.60–0.20): Absent perfusion A positive CAD diagnosis required ≥1 segment with stress-induced TPR <0.97, aligning with ESC/ACC 2024 guidelines [17, 18]. MDCT perfusion served as the reference standard. Post-procedurally, all patients received cardiology consultations for evidence-based treatment planning.

### Statistical analysis

Distribution normality for quantitative parameters was assessed using the Shapiro-Wilk test. Descriptive statistics included mean ± standard deviation (SD), median with interquartile range (IQR), and minimum/maximum values. Categorical features were summarized as absolute frequencies (n) and proportions (%).

Comparative analyses employed Welch’s t-test for normally distributed quantitative data (two-group comparisons) and the Mann-Whitney U-test for non-normally distributed data. Categorical variable comparisons used Pearson’s chi-square test, with Fisher’s exact test applied where chi-square assumptions were violated.

Statistical processing carried out using the R programming language v4.2, Python v.3.10 [^R], Statistica 12 programme. (StatSoft, Inc. (2014). STATISTICA (data analysis software system), version 12. www.statsoft.com.), and SPSS IBM version 28. P considered statistically significant at <0.05.

### Method of machine learning model building with cross-validation

The study developed a machine learning model for non-invasive IHD diagnosis using resting ECG parameters validated against myocardial perfusion imaging as the gold standard. Data integration combined two primary sources: (1) a primary dataset containing gold-standard labels of myocardial perfusion defects post-stress ATP, and (2) resting ECG measurements from a Cardio-Qvark system. Preprocessing standardized labels into binary classes (IHD=1, healthy=0), addressed ambiguous entries through exclusion, and handled missing values via feature-wise median imputation. All features were standardized to zero mean and unit variance using StandardScaler to ensure comparability across parameters.

For model development, we implemented two interpretable algorithms: a Random Forest (RF) with 100 estimators, maximum depth constrained to 3, and balanced class weighting to mitigate bias; and a Lasso-regularized Logistic Regression (LR) with liblinear solver and balanced class weighting. Model selection employed stratified 5-fold cross-validation—preserving class distribution in each split—with AUC-ROC as the primary performance metric. The best-performing model was selected based on superior mean cross-validated AUC and subsequently retrained on the full dataset.

Performance evaluation incorporated both discriminative and clinical diagnostic metrics. We reported AUC-ROC, accuracy, sensitivity, specificity, F1-score, positive predictive value (PPV), and negative predictive value (NPV). To quantify uncertainty, bootstrap resampling (1,000 iterations) generated 95% confidence intervals using the percentile method for all primary metrics. Model interpretability was enhanced through feature importance analysis, identifying the top 20 predictive ECG parameters using either Gini importance (RF) or absolute coefficient magnitudes (LR).

Methodological rigor addressed key challenges: class imbalance was mitigated via stratified sampling and algorithm-level class weighting; overfitting was controlled through L1 regularization (LR) and depth constraints (RF); and reproducibility was ensured by fixing random seeds. The pipeline automatically generated a comprehensive diagnostic report including dataset characteristics, cross-validation results, classification metrics, confusion matrix, and bootstrap-derived confidence intervals—providing clinically actionable metrics with uncertainty estimates.

## Results

The descriptive features of the study participants described in the table below. (Table 1 and Table 2)

**Table 1.** Comparative characteristic of the sample and groups.

| Index | Whole sample mean ± SD | 1<sup>st</sup> group mean ± SD | 2<sup>nd</sup> group mean ± SD | P |
| --- | --- | --- | --- | --- |
| Age (years) | 56.28 ± 10.60 | 59.93 ± 11.70 | 53.96 ± 9.23 | 0.013* |
| Pre-test probability of IHD (%) | 8.81 ± 7.71 | 9.90 ± 8.14 | 8.12 ± 7.42 | 0.31 |
| HR (beat/mint) | 70.29 ± 9.55 | 70.22 ± 10.74 | 70.32 ± 8.84 | 0.96 |
| SBP at rest (mmHg) | 123.16 ± 15.43 | 124.48 ± 20.56 | 122.32 ± 11.22 | 0.54 |
| DBP at rest (mmHg) | 80.61 ± 11.23 | 82.35 ± 13.16 | 79.51 ± 9.81 | 0.27 |
| Weight (kg) | 77.92 ± 16.23 | 77.12 ± 14.71 | 78.42 ± 17.25 | 0.73 |
| height (cm) | 169.95 ± 8.83 | 169.80 ± 9.41 | 170.04 ± 8.54 | 0.90 |
| BMI (kg/m<sup>2</sup>) | 26.93 ± 4.90 | 26.70 ± 4.23 | 27.06 ± 5.31 | 0.75 |
| Heart rate at rest (Cardio-Qvark®) (beat/mint) | 69.68 ± 9.26 | 69.64 ± 11.51 | 69.69 ± 7.63 | 0.98 |
| CardioQVARK vessel elasticity (%) | 23.012 ± 24.36 | 20.96 ± 22.41 | 24.33 ± 25.68 | 0.55 |
| Ejection fraction (%) | 64.52 ± 4.38 | 64.47 ± 4.77 | 64.54 ± 4.22 | 0.95 |
| Creatinine (μmol/L) | 82.74 ± 16.01 | 80.37 ± 15.44 | 84.23 ± 16.34 | 0.29 |
| eGFR | 85.31 ± 14.68 | 84.85 ± 15.10 | 85.58 ± 14.53 | 0.82 |
| Statistical significance was defined as $p < 0.05$ | | | | |
| Continuous variables are reported as mean ± standard deviation (SD). | | | | |

| **Index** | **Whole sample mean ± SD** | **1<sup>st</sup> group mean ± SD** | **2<sup>nd</sup> group mean ± SD** | **P** |
| --- | --- | --- | --- | --- |
| Independent samples were compared using Student's t-test.<br>Abbreviations: systolic blood pressure (SBP), diastolic blood pressure (DBP), body mass index (BMI), heart rate (HR), estimated glomerular filtration rate (eGFR), and ischemic heart disease (IHD) |  |  |  |  |

**Table 2.**
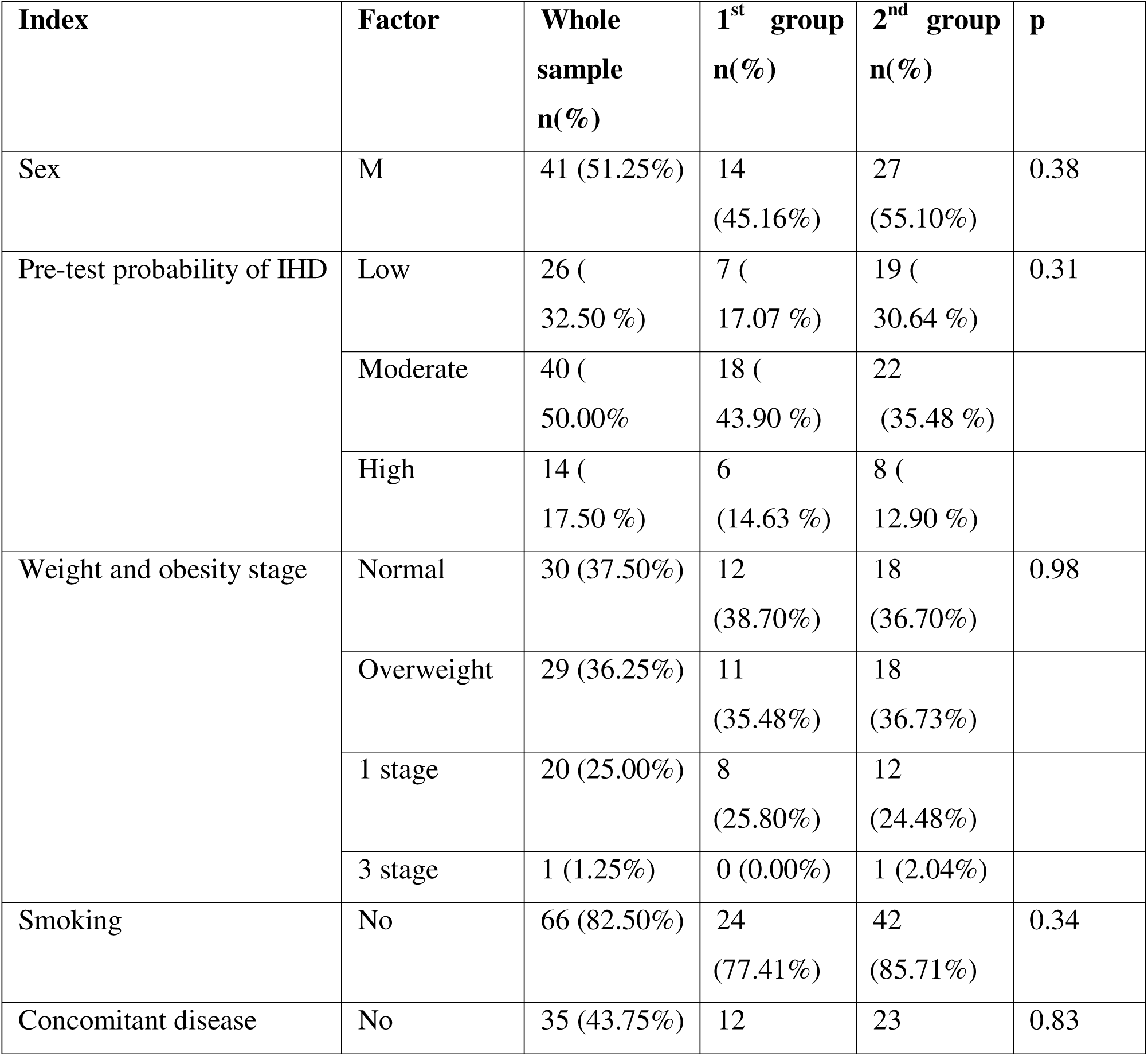
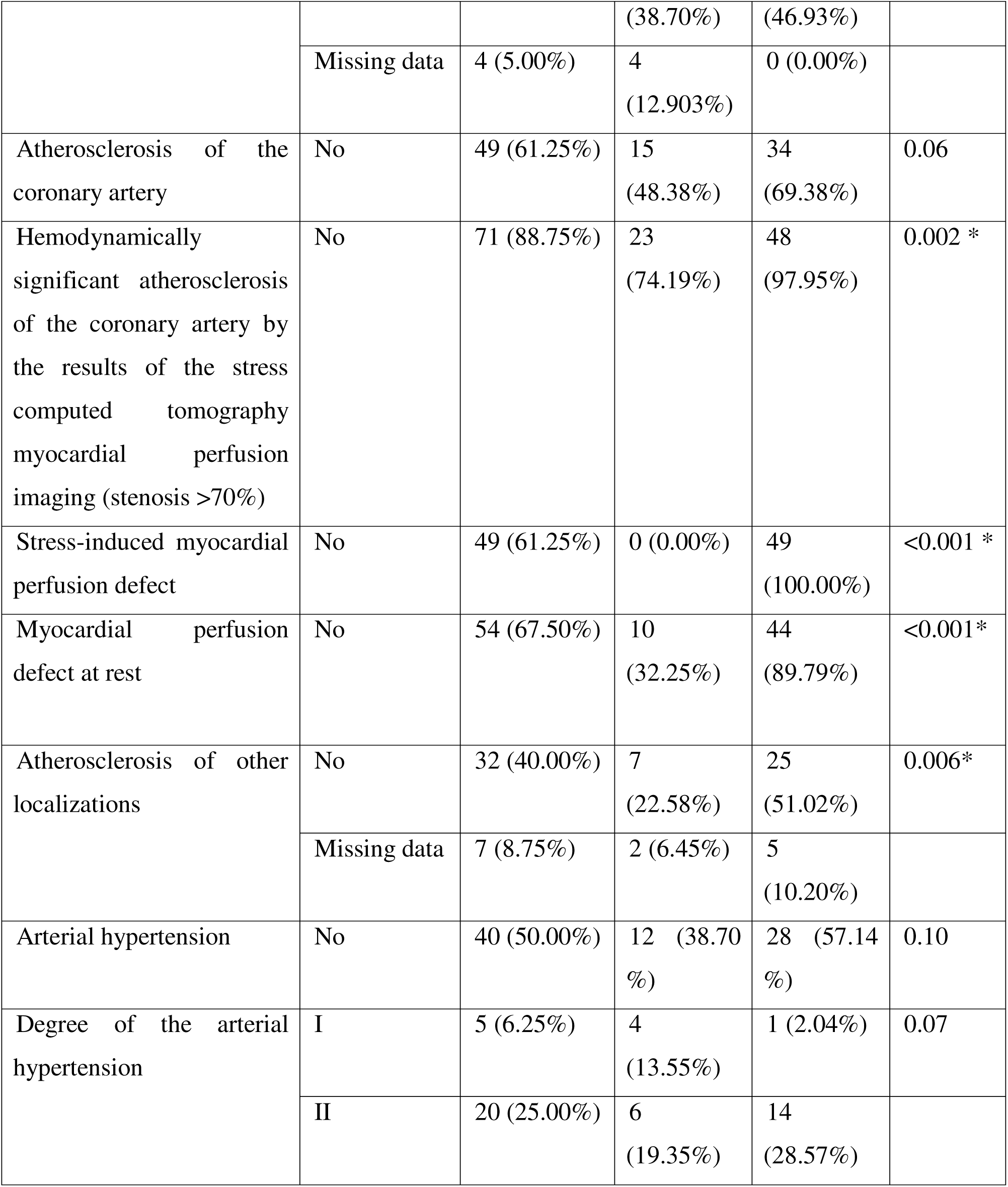
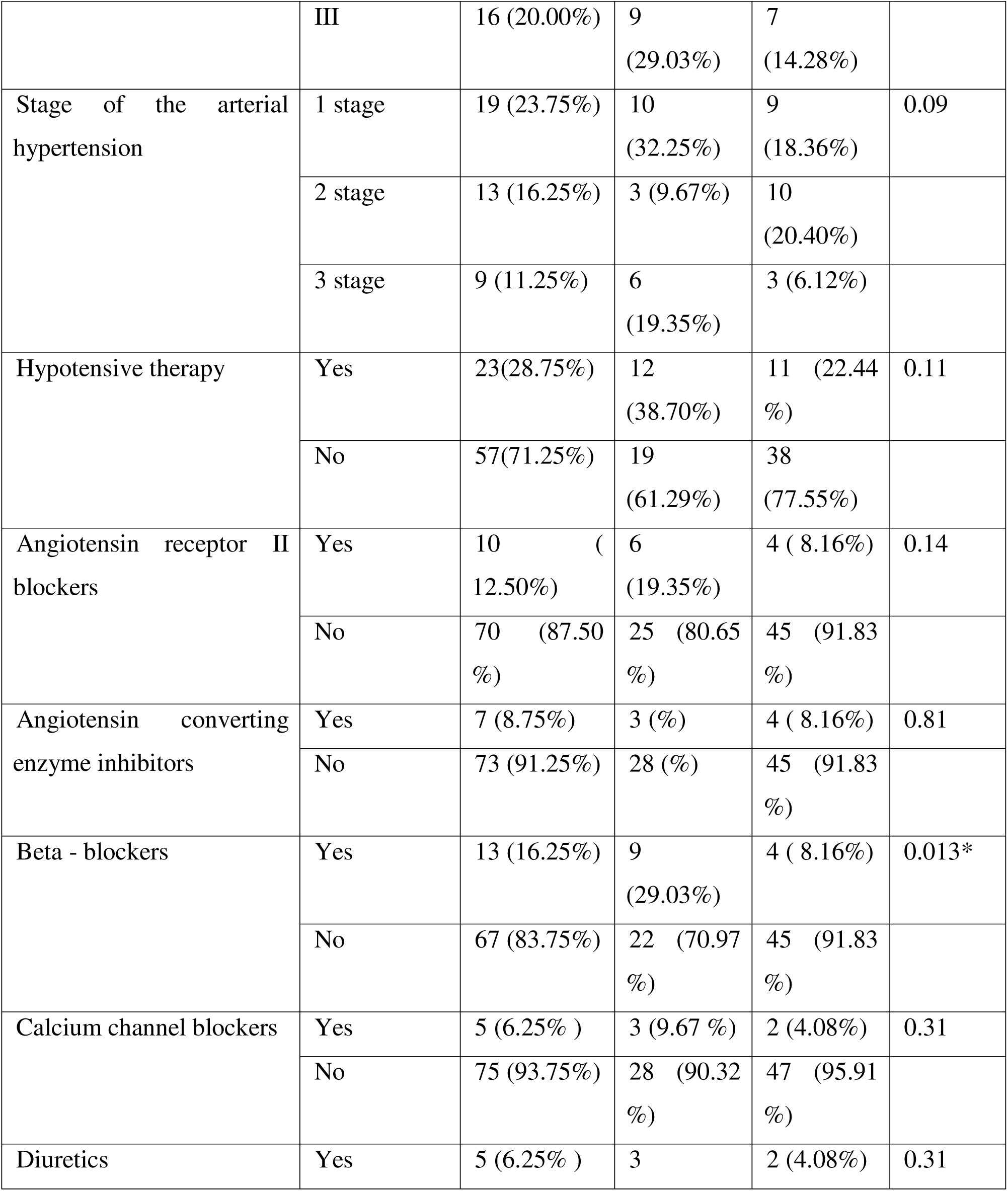
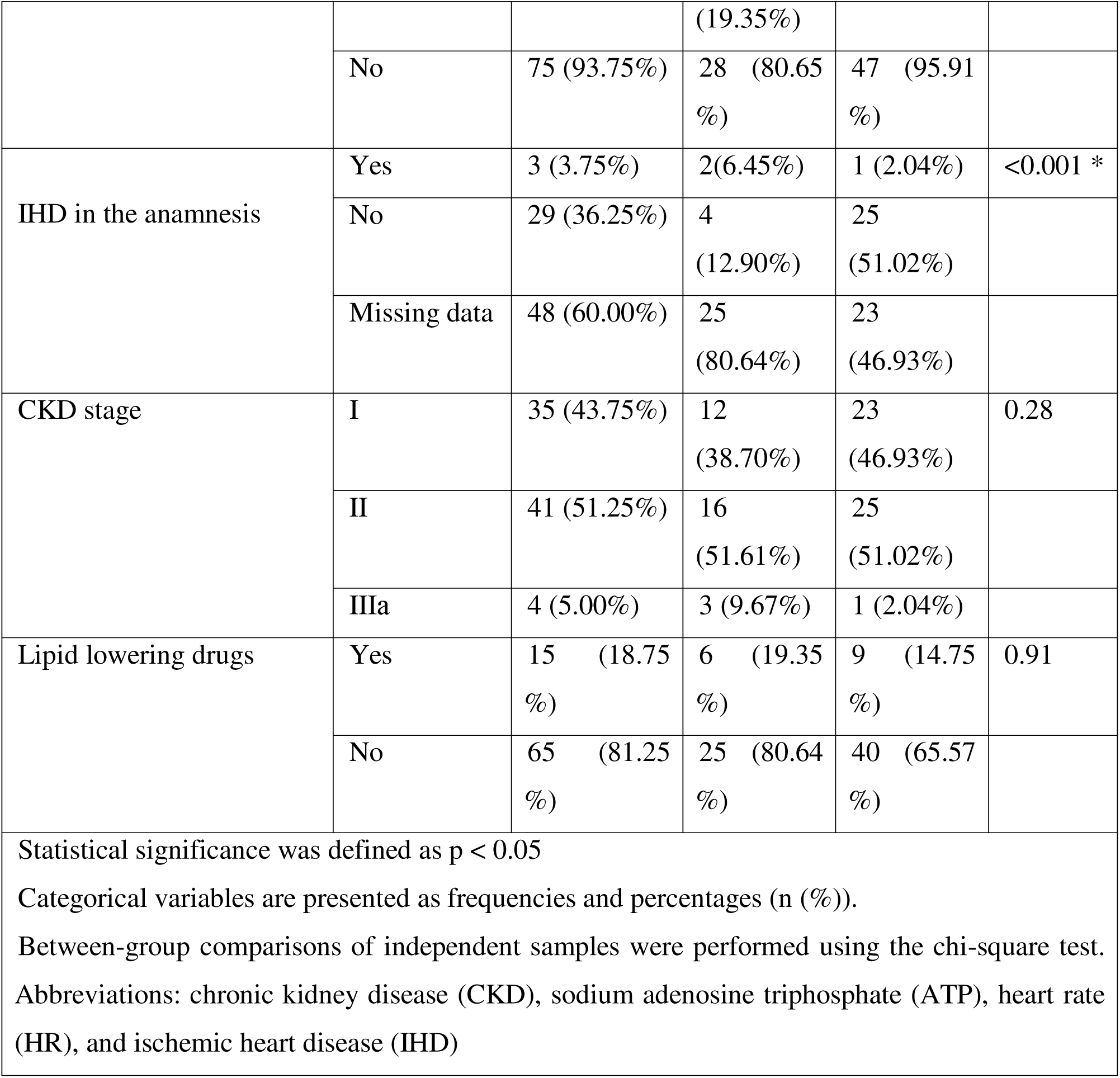
A comparative analysis of categorical variables of the sample and the groups.

### The built Random Forest machine learning model performance in the diagnosis of IHD

According to the methods explained in the section “material and methods”, the built machine learning performance in the diagnosis of ischemic heart disease represented in the figure 1 and table 3 below.

**Figure 1.** The diagnostic performance of the Random Forest machine learning model in the diagnosis of ischemic heart disease based on the single lead electrocardiography parameters.

**Table 3.** The diagnostic performance of the Random Forest machine learning model in the diagnosis of ischemic heart disease based on the single lead electrocardiography parameters.

| Metrics | Mean value | 95 % confidence interval |
| --- | --- | --- |
| AUC | 0.988 | [0.967-1.000] |
| Sensitivity | 0.871 | [0.739-0.971] |
| Specificity | 0.959 | [0.894-1.000] |
| Positive Predictive Value | 0.931 | [0.833-1.000] |
| Negative Predictive Value | 0.922 | [0.837-0.982] |

The following single lead electrocardiography parameters are the mathematically most important for the diagnosis of ischemic heart disease based on the built Random Forest machine learning model. (Table 4)

**Table 4.** The top 20 features (parameters) of the single lead electrocardiography (ECG) in the diagnosis of ischemic heart disease according to the Random Forest machine learning model. These parameters are mathematical calculations calculated automatically when registering the single lead ECG on the Cadio-Qvark® device.

| Feature | Importance coefficient |
| --- | --- |
| Sbeta | 0.049990 |
| Tfi | 0.046833 |
| Beta | 0.046738 |
| QTc | 0.046383 |
| PpeakN | 0.035806 |
| Pan | 0.034780 |
| PpeakP | 0.031999 |
| J80A | 0.031847 |
| Tons | 0.027750 |
| QRSfi | 0.027592 |
| SDNN | 0.026740 |
| Rpeak | 0.026401 |
| SA | 0.025739 |
| HFNoise | 0.024630 |
| Tpeak | 0.024480 |
| QRS11energy | 0.023391 |
| Speak | 0.022925 |
| Pfi | 0.022143 |
| Pst | 0.022038 |
| TA | 0.021383 |

## Discussion

This study presents compelling evidence for the high diagnostic accuracy of a machine learning (ML) model, specifically a Random Forest (RF) algorithm, utilizing resting single-lead electrocardiography (SLECG) parameters derived from the portable Cardio-Qvark® device in the non-invasive detection of ischemic heart disease (IHD), validated against stress myocardial perfusion imaging (MPI) as the gold standard. The exceptional performance metrics, notably an AUC of 0.988 (95% CI: 0.967-1.000), sensitivity of 0.871, and specificity of 0.959, suggest that this approach holds significant promise for transforming IHD screening and diagnosis.

The primary strength of this research lies in the demonstration that sophisticated analysis of simple, rapidly acquired, and entirely non-invasive resting SLECG signals can achieve diagnostic accuracy approaching that of more complex, expensive, and potentially risky tests like stress MPI. This finding is particularly relevant given the critical need for accessible, cost-effective, and scalable tools for early IHD detection, especially in primary care settings, resource-limited environments, or for large-scale screening programs. The Cardio-Qvark® device represents a practical platform for this application, enabling data collection outside traditional clinical settings. The identification of mathematically significant ECG parameters (e.g., Sbeta, Tfi, Beta, QTc, PpeakN) by the RF model provides insights into the electrophysiological features most discriminatory for underlying ischemia. While the exact physiological interpretation of some of these algorithmically derived features requires further investigation, their association aligns with established ECG markers of ischemia (e.g., QT interval prolongation, T-wave changes) and autonomic function (e.g., heart rate variability like SDNN). The model’s focus on parameters like HFNoise also suggests it effectively incorporates signal quality assessment.

The methodological rigor employed in model development enhances the credibility of these results. The use of stratified 5-fold cross-validation mitigates overfitting and provides a robust estimate of generalizability. Addressing class imbalance through balanced class weighting within the RF algorithm was crucial given the group sizes (n=31 defect vs. n=49 non-defect). The inclusion of bootstrap-derived 95% confidence intervals for all primary performance metrics provides a clear understanding of the precision of the estimates. Furthermore, the choice of interpretable ML models (RF with constrained depth and Lasso Logistic Regression) and the reporting of top features enhance the clinical translatability and potential for understanding the model’s decision-making process compared to “black-box” alternatives.

The comparative analysis of baseline characteristics (Tables 1 & 2) provides essential context. The significant difference in age between groups (Group 1 older, p=0.013) aligns with the known epidemiology of increasing IHD risk with age. Crucially, the groups showed no significant differences in most traditional risk factors like resting heart rate, blood pressure, BMI, smoking status, or eGFR. However, the expected and highly significant differences were observed in the key pathological endpoints defining the groups: presence of stress-induced perfusion defects (p<0.001), rest perfusion defects (p<0.001), and hemodynamically significant stenosis (>70%, p=0.002). Group 1 also had a significantly higher prevalence of atherosclerosis in other vascular beds (p=0.006), beta-blocker use (p=0.013), and prior IHD history (p<0.001), all consistent with a cohort experiencing more advanced cardiovascular disease. The lack of significant differences in many baseline variables strengthens the argument that the ML model’s discriminatory power stems primarily from its analysis of the ECG signal itself rather than confounding demographic or clinical factors.

The implications of these findings are substantial. A highly accurate, non-invasive, low-cost, and rapid screening tool for IHD based on a simple ECG recording could revolutionize patient pathways. It could serve as a powerful triage tool in primary care or emergency departments, potentially reducing unnecessary referrals for more expensive and invasive tests like coronary angiography in low-risk individuals identified by the model as healthy. Conversely, its high sensitivity could help identify individuals warranting prompt further investigation with gold-standard modalities like MPI or CT coronary angiography. The portability of the Cardio-Qvark® device further extends its potential utility to point-of-care settings, remote monitoring, or community-based screening initiatives. The model’s reliance on resting ECG also circumvents the contraindications and risks associated with stress testing.

Despite the highly promising results, several limitations must be acknowledged. Firstly, the sample size (n=80) is relatively modest. While sufficient for the primary ML analysis given the strong effect size, larger, multi-center validation studies are essential to confirm generalizability across diverse populations and healthcare settings. Secondly, the single-center design at Sechenov University introduces potential selection bias and limits the external validity of the findings. Thirdly, the retrospective inclusion of healthy controls, while pragmatic, introduces a methodological asymmetry compared to the prospective enrolment of the pathology group. Future studies should prospectively enroll both cases and controls using identical protocols. Fourthly, while validated against MPI, the study did not correlate findings with invasive coronary angiography in all participants, which remains the definitive anatomical standard for coronary stenosis. Further research correlating the ECG-derived features and model predictions directly with angiographic findings would be valuable. Finally, the model’s performance in specific subpopulations (e.g., patients with bundle branch blocks, arrhythmias, or paced rhythms) remains unexplored and warrants investigation.

The pathophysiological link between the identified top ECG features and myocardial ischemia requires deeper exploration. While some features (QTc, SDNN, Tpeak) have established links to repolarization abnormalities and autonomic dysfunction associated with IHD, others (Sbeta, Tfi, Beta, PpeakN) represent more complex mathematical derivations. Future research should focus on elucidating the biological underpinnings of these features to enhance clinical understanding and potentially identify novel electrophysiological markers of ischemia. Additionally, exploring the model’s ability to quantify ischemia severity or localize affected coronary territories would significantly enhance its clinical utility beyond binary classification.

Previous study on the use of the single lead parameters ECG in the diagnosis of ischemic heart disease showed a relatively low diagnostic accuracy, AUC 67 % [9].

### Conclusion

In conclusion, this study demonstrates the exceptional diagnostic accuracy of a machine learning model utilizing resting single-lead ECG parameters acquired via the Cardio-Qvark® device for detecting IHD, as defined by stress-induced myocardial perfusion defects on MPI. The methodological rigor, combined with the compelling performance metrics (AUC 0.988), positions this approach as a highly promising, non-invasive screening tool. Its simplicity, portability, and potential cost-effectiveness make it particularly attractive for widespread implementation. While larger, prospective, multi-center validation studies are crucial next steps, these findings represent a significant advancement towards accessible and accurate point-of-care detection of ischemic heart disease, paving the way for earlier intervention and improved patient outcomes. Future research should focus on validating these results in broader populations, elucidating the pathophysiology behind key features, and integrating the tool into real-world clinical decision-making pathways.

## Decelerations

1. Ethics approval and consent to participate: the study approved by the Sechenov University, Russia, from “Ethics Committee Requirement № 19-23 from 26.10.2023”. An informed written consent is taken from the study participants.
2. Consent for publication: applicable on reasonable request
3. Availability of data and materials: applicable on reasonable request
4. Competing interests: The authors declare that they have no competing interests regarding publication.
5. Funding’s: The work of Philipp Kopylov was financed by the government assignment 1023022600020-6 «Application of mass spectrometry and exhaled air emission spectrometry for cardiovascular risk stratification». The Work of Philipp Kopylov was financed by the Priority 2030 program of the Ministry of Science and Higher Education of Russia, project “Screening of cardiac pathology using telemedicine technologies and elements of artificial intelligence”, code 03.000.B.163. The work of Basheer Marzoog was financed by the Priority 2030 program of the Ministry of Science and Higher Education of Russia, project «The Digital Cardiology with Artificial Intelligence».
6. Authors’ contributions: MB is the writer, researcher, collected and analyzed data, interpreted the results. and revised the final version of the paper, biostatistical analysis of the sample and PhK revised the final version of the manuscript. All authors have read and approved the manuscript.
7. The paper has not been submitted elsewhere
8. Declaration of AI use: not used

## Data Availability

All data produced in the present work are contained in the manuscript

## Acknowledgments

Not applicable

## Authors’ information

**Basheer Abdullah Marzoog**, Institute of Personalized Cardiology of The Center “Digital Biodesign and Personalized Healthcare” of Biomedical Science and Technology Park, Federal State Autonomous Educational Institution of Higher Education I.M. Sechenov First Moscow State Medical University of the Ministry of Health of the Russian Federation (Sechenovskiy University), 119991 Moscow, Russia; postal address: Russia, Moscow, 8-2 Trubetskaya street, 119991. **Philipp Kopylov,** director of the Institute of Personalized Cardiology of The Center “Digital Biodesign and Personalized Healthcare” of Biomedical Science and Technology Park, Federal State Autonomous Educational Institution of Higher Education I.M. Sechenov First Moscow State Medical University of the Ministry of Health of the Russian Federation (Sechenovskiy University),119991 Moscow, Russia; postal address: Russia. Moscow, 8-2 Trubetskaya street, 119991. ORCID: 0000-0002-4535-8685. Scopus ID: 6507736224.

## STANDARDS OF REPORTING

– STROBE guideline has been followed.
– The TRIPOD+AI standard of reporting for prediction models has been followed

## Key Messages

- A machine learning model analyzing resting single-lead ECG signals achieves exceptional accuracy (AUC 0.988) in diagnosing ischemic heart disease, validated against stress myocardial perfusion imaging.
- This approach transforms a simple, portable ECG device into a potent, non-invasive screening tool, offering a practical and accessible alternative to complex, costly diagnostic tests.
- The findings pave the way for point-of-care IHD detection, enabling earlier diagnosis, optimized resource allocation, and improved patient outcomes, particularly in primary care and resource-limited settings.

